# Generation of machine-learning derived Cancer Vulnerability Indicator to determine the spatial burden of cancer outcomes

**DOI:** 10.1101/2025.02.06.25321783

**Authors:** Kou Kou, Jessica Cameron, Paramita Dasgupta, Hao Chen, Peter D Baade

## Abstract

**Background:** Due to the difficulty of obtaining population-based individual-level data, ecological studies are often used to explore factors related to geographic variations in health outcomes. This study proposes a novel framework to identify area-level predictors of spatial variations in lung cancer outcomes and generate a lung cancer vulnerability index (LcVI) based on these predictors.

**Methods:** Data on 11,313 persons diagnosed with invasive lung cancer in Queensland, Australia (2016-2019) were sourced from the population-based Queensland Cancer Register. Bayesian spatial models estimated smoothed standardised incidence ratios (SIRs) for 519 geographic areas. Area-level variables (n= 911) were extracted from multiple data collections. Random forest models were fitted to identify important predictors for lung cancer incidence rates. A novel non-parametric dimensionality reduction approach incorporating the final random forest model results was developed to generate the LcVI which ranged from 0-10.

**Results:** Eight variables were identified as predictors for lung cancer incidence with the top two being the prevalence of diabetes and adequate fruit intake. Areas having incidence rates below the Queensland average had significantly lower LcVI than those with average incidence rates (mean difference = 2.80, 95% CI: 2.34-3.25, p < 0.001) while areas with above average incidence rates had significantly higher LcVI than those with average incidence (mean difference = 2.70, 95% CI: 2.20-3.19, p < 0.001). The LcVI was strongly associated with the continuous SIR, explaining 57% of the variation (R² = 0.57, p< 0.001).

**Conclusion:** This novel approach identified a small number of important predictors for lung cancer incidence from a high-dimensional dataset. The lung cancer vulnerability index based on these predictors effectively explained the geographic variations in incidence, potentially offering insights into the underlying drivers of these variations. The favourable performance of this approach may promote further ecological studies on other cancer outcomes.

## Introduction

With the expansion of cancer registries worldwide, we now have the ability to map geographic patterns of cancer outcomes [1, 2], presenting a valuable opportunity to investigate the factors driving these variations. Since the data describing the potential drivers can be difficult to obtain at the individual level across the whole population, ecological studies using aggregated socio-environmental variables are commonly used to reveal important associations [3]. However, the vast amount of area-level data that are currently available poses challenges for traditional statistical models, which can struggle to manage the complexity of high-dimensional datasets.

Lung cancer was the most frequently diagnosed cancer and the leading cause of cancer-related deaths, globally responsible for almost 2.5 million new cases in 2022 [1]. Based on the Australian Cancer Atlas 2.0 [2], there was a four-fold disparity in small area-specific lung cancer incidence rates across Australia. Understanding the factors relating to this geographic disparity is crucial for the distribution of health resources and disease burden projection, especially as lung cancer is typically diagnosed at an advanced stage [4]. However, many studies that have examined the relationships between area-level socioeconomic indices and lung cancer incidence have reported inconsistent findings [1, 5–7]. This inconsistency may be partly due to differences in the methodologies used to construct such indices [8], which are often generated using diverse socio-environmental variables without fully considering their direct relevance to cancer outcomes. Additionally, many dimensionality reduction methods that are commonly used for wide data sets, methods such as factor analysis or principal component analysis (PCA), combine correlated variables into artificial features, without assigning weights that reflect the specific associations of variables with cancer outcomes [9].

In this study, we developed a machine learning-based approach to identify important predictors of geographic patterns in cancer outcomes using routinely collected area-level data. Key predictors were then combined using a novel dimensionality reduction method, to generate a cancer vulnerability index (CVI) that helped explain spatial variations in cancer outcomes. This approach was applied to population-based lung cancer incidence data in Queensland, Australia.

## Methods

Approval was obtained from the data custodian to access de-identified routinely collected cancer incidence data from Queensland Cancer Register (QCR). The QCR is a legislated population-based registry. It is required by law for information on all invasive cancers (excluding keratinocyte cancers) diagnosed in Queensland to be notified to the QCR [10]. Access to the QCR de-identified data extract for research purposes was provided on September 2, 2024 and the authors did not have access to information that could identify individual participants. Data quality is high, with 93% of cases histologically verified and 0.9% diagnosed by death certificate only in 2016 (unpublished data, QCR).

### Outcome of interest (Figure 1, Orange boxes)

Records for all persons (n = 11,313) diagnosed with a primary invasive bronchial or lung cancer (ICD-10 code: C34.0-34.9) aged 20 or over in Queensland from 2016-2019 were extracted from the QCR. Cases without residential information (n=39) and those with multiple primary lung cancer diagnoses (n=67) were excluded, giving a final study cohort of 11,207 individuals. Patients’ residential information at lung cancer diagnosis was defined by the 2016 Statistical Area Level 2 (SA2) [11]. In 2016 there were 528 SA2s covering Queensland without gaps or overlaps, of varying land area (median area 14 km^2^, interquartile range (IQR): 6 to 95 km^2^) and population (median: 7,857, IQR: 4,922 to 11,331) [11]. Nine SA2s with small populations (<5 residents annually, on average) were excluded from statistical modelling, leaving 519 SA2s for analysis.

**Figure 1.**
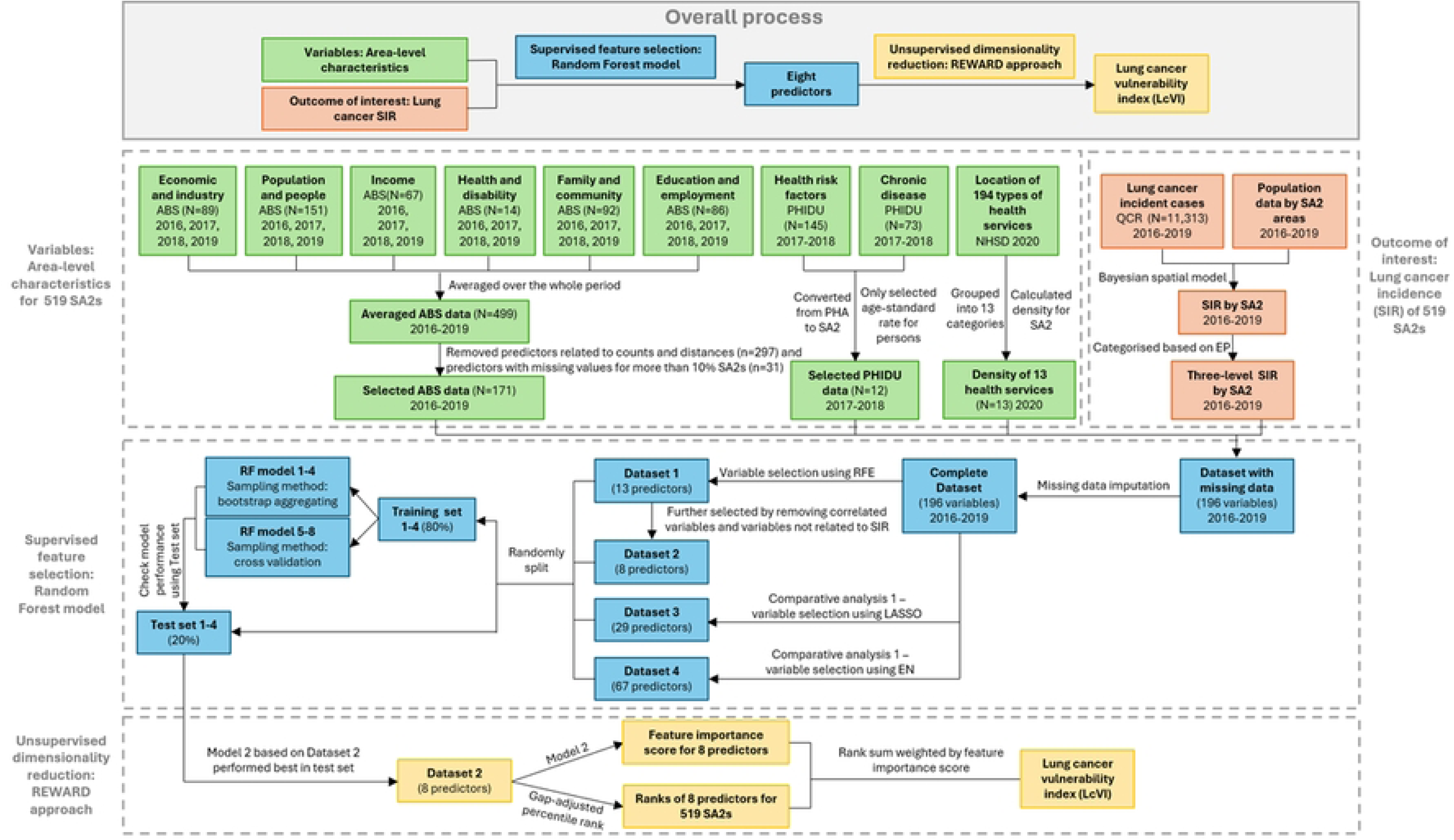
Flowchart of generating machine learning derived lung cancer vulnerability index (LcVI) Note: SIR: Standardised incidence ratio; ABS: Australian Bureau of Statistics; PHIDU: Public Health Information Development Unit; NHSD: National Health Services Directory; QCR: Queensland Cancer Register; PHA: Population Health Area; SA2: Statistical Area Level 2; SIR: Standardized incidence ratio; PP: Posterior probability; RFE: Recursive feature elimination; LASSO: Least absolute shrinkage and selection operator; EN: Elastic net; RF: Random forest.

The outcome of interest was the smoothed indirect standardized incidence ratios (SIR) of lung cancer during 2016-2019. SIRs reflect the relative incidence rate of lung cancer in each SA2 compared to the Queensland average, with high SIR values (>1) indicating that the incidence was above the Queensland average; SIR=1 indicating similar incidence to the average; and low SIR values (<1) indicating below average incidence. The methods for estimating the SIRs using a Bayesian spatial model have been reported elsewhere [12, 13] and further details are in Appendix 1.

In addition, the exceedance probability (EP) for each SA2 was estimated from the SIR model, and these provide statistical evidence indicating whether an area’s incidence rates are greater than the state average [14]. Based on previous studies [15], we classified SA2s into three categories based on EP values, with low EP values (<0.2) providing evidence that incidence rates in those SA2s were below average (referred to as “below average”); values between 0.2 and 0.8 suggested a lack of evidence for a difference from the Queensland average (“average”); and high EP (>0.8) indicated the incidence rates were above average (“above average”).

Population data for each SA2 by sex, calendar year and five-year age groups were obtained from the Australian Bureau of Statistics (ABS) [16].

### Area-level characteristics (Figure 1, green boxes)

Area-level data from the ABS, Public Health Information Development Unit (PHIDU), and National Health Services Directory (NHSD) were retrieved from the Australian Urban Research Infrastructure Network (AURIN) Data Provider [17].

The ABS-sourced area level data for 2011-2019 [18] included 89 economic and industry [19], 151 population and people [20], 67 income [21], 14 health and disability [22], 92 family and community [23], and 86 education and employment variables [24]. For each SA2, data for each variable for at least one of the single years of 2016 to 2019 were extracted and averaged over the number of years the variable was available. Variables with missing average values for more than 10% of the 528 SA2s were excluded (n=31). In addition, variables related to counts which are not comparable across SA2s were also excluded (n= 297), leaving 171 variables for analysis (Appendix 2).

The PHIDU-sourced [25] dataset included 145 variables relating to adult risk factors [26] and 73 variables relating to chronic disease in 2017-2018 [27]. Originally available at the level of Population Health Areas (PHA) which comprised a mix of single and aggregated SA2 areas, these were converted to SA2 via a population-weighed correspondence file [28]. Only variables reporting the age standardised rates (ASR) of each chronic disease and risk factor for persons were selected (n=12, Appendix 2).

The NHSD [29] provided information on 194 types of health services by longitude and latitude coordinates, which were grouped into 13 categories (Appendix 3&4). In Queensland, as of November 2020, a total of 24,693 facilities fell within these 13 categories. For each SA2, the density per square kilometre for each category was determined based on the number of services within a buffered region of the SA2, determined by the SA2’s geographical size and a 10-kilometer traversable accessibility network distance (as described in Appendix 5).

The final combined dataset included 196 area-level variables for each SA2 for 2016-2019.

### Random Forest Weighted Gap Adjusted Percentile Rank Sum Index (REWIRED)

The REWIRED approach can be subdivided into two principal sections, a supervised variable selection phase (Figure 1, Blue boxes), and an unsupervised dimensionality reduction phase (Figure 1, Yellow boxes).

#### Supervised feature selection (Figure 1, Blue boxes)

The input data for supervised variable selection was the three-category outcome for lung cancer incidence (“below average”, “average”, “above average”) and the 196 area-level variables for the 519 SA2s. This process incorporated missing data imputation, feature selection, model training, model performance evaluation, and feature importance estimation.

A random forest approach was used for imputation, and model building. Random forest is a non-parametric classification and regression tool for constructing prediction rules, that combines multiple tree predictors, with each tree trained on a sample randomly drawn from the original dataset using different sampling methods [30]. One advantage of random forests is that they do not make any prior assumptions on the form of their association with the outcome variable [30].

In the dataset, 106 out of 519 (20.4%) SA2s had at least one missing value among the 196 area-level variables. Missing data for the variables were imputed using a random forest model with the bagging (bootstrap aggregating) sampling method [31]. Bagging creates multiple bootstrap samples from the original dataset with replacement, and is robust against noisy data and outliers [32]. For missing data imputation, a decision tree is trained on each bootstrap sample using the remaining variables as predictors to predict the missing values. The final imputed value for each missing data point was obtained by averaging the predictions from all the trees. The random forest method has been demonstrated to significantly improve accuracy in missing data imputation compared to parametric imputation models [33]. After imputation, the 196 variables were centred by subtracting the mean value of each variable and scaled by dividing by its standard deviation.

We applied a non-parametric feature selection method, recursive feature elimination (RFE), to select predictor variables for the final random forest model [34]. RFE uses the random forest model to evaluate the importance of each predictor iteratively and eliminate the least important predictors in a backward manner. To increase the robustness of the final index [35], we set the RFE to select no more than 15 predictors, and this set of predictors was labelled the “RFE predictors”. In addition, we identified a smaller subset of predictors by removing any predictors that had a correlation coefficient over 0.7 or less than −0.7 with other more important predictors; and any predictors that were not significantly associated with the continuous log SIR in linear regressions, and this set was labelled the “RFE_selected predictors”. The model performances using these two sets of variables were compared using a 80% training set and 20% test set, ensuring that the distribution of the outcome variable was maintained in both sets.

Random forest involves several key hyperparameters including the number of decision trees in the forest; the minimal number of observation in a node (nodesize); and the number of predictors drawn randomly from the predictor list to include as candidate splitting predictors at each split (mtry) [36]. The number of trees is determined by the number of independent sub-samples drawn from the dataset, which depends on the sampling method. We compared two commonly used sampling methods in the model training process: bagging and K-fold cross-validation (CV). K-fold cross-validation is a method that divides the training dataset into K equal sets of observations, where each part is used once as validation data while the remaining K-1 parts are used for training [37, 38]. This process is repeated K times, to train the random forest model, allowing for a more reliable estimate of its performance by averaging results across all K folds. To develop the final random forest models, we used bagging with 100 bootstrap samples for models 1 and 2; and 10-fold CV for model 3 and 4. We selected nodesize and mtry values based on the ones that yielded the highest Kappa, a model performance statistic (Appendix 6) [39].

The performance of the models was tested on the test dataset and accuracy was assessed using confusion matrices (Appendix 6) [40]. The model with the highest accuracy and Kappa value was chosen as the final model. Feature importance scores of the predictors were extracted from the results of this final model.

#### Unsupervised dimensionality reduction (Figure 1, Yellow boxes)

In this phase, the lung cancer vulnerability index (LcVI) was generated using the selected predictors and their feature importance scores via the REWIRED approach. This approach first rescaled the values for each predictor using the ‘gap adjusted percentile rank’ (described below), then weighted the rescaled values using their feature importance scores, and finally summed the weighted values for each area to create the LcVI.

The conventional percentile ranking method relies on the sequences of values, overlooking the absolute magnitude of differences between adjacent values. To address this limitation, we developed a ‘gap-adjusted percentile rank,’ accounting for varying intervals between values.

This approach identified the minimum and maximum values of each continuous predictor, rescaling the predictor into 100 levels. Each observation was then assigned a rank based on its position within these levels, allowing for a standardized comparison within the predictor (Appendix 7). The gap-adjusted percentile rank (*p*_*va*_) for a specific area (*a*) and predictor (*v*_*a*_) can be calculated as:

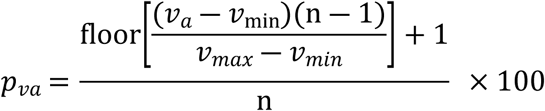

Where *v*_*max*_ is the maximum value for variable v; *v*_*min*_ is the minimum value for variable v; n is the number of observations; 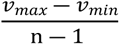 is the uniform gap between neighbouring ranks; floor 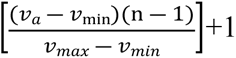 is the gap adjusted rank (‘floor’ rounds down to the nearest integer; ‘+ 1’ ensures ranks start at 1).

Following the ‘gap-adjusted percentile rank’ approach, predictor values were rescaled in ascending order. For predictors negatively associated with the outcome, values were inversely rescaled to ensure that the gap-adjusted percentile values for all predictors maintained a positive association with the outcome.

In contrast to the conventional percentile rank-sum approach, which merely aggregates percentile values of a set of variables, our approach also employed weighted summation to construct the LcVI. The weights of the predictors were the feature importance scores estimated from the final Random Forest model. The formulation is expressed as:

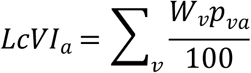

Where *LcVI*_*a*_ is the Lung cancer Vulnerability Indicator for area a; *W*_*v*_ is the feature importance scores for predictor v; *p*_*va*_ is the gap-adjusted percentile rank for predictor v in area a.

Extreme LcVI values below the 5th percentile and above the 95th percentile were truncated at these thresholds. Values were centred by subtracting the truncated minimum value and scaled by dividing the truncated range and then multiplying by 10 to get the final LcVI index values ranging from zero to ten. Hence zero represented the least vulnerable and ten the most vulnerable areas for the diagnosis of lung cancer.

An ANOVA test followed by a post-hoc analysis using Tukey’s Honest Significant Difference (HSD) test, a linear regression model, and various visualisations were used to evaluate the capacity of the LcVI to explain the geographic variation of lung cancer incidence in Queensland.

### Comparative analysis

Parametric variable selection methods were also used to select predictors and compare estimated models with the RFE models (Figure 1, Blue boxes). We applied Least Absolute Shrinkage and Selection Operator (LASSO) regression and Elastic Net (EN) regression as the parametric feature selection methods [41, 42] (Appendix 8). The performance of models 5 to 8 based on features selected using LASSO and EN were compared with models 1 to 4 (Table 1).

**Table 1.** Model performance based on test set.

| Model<sup>1</sup> | No.<br>predictors<sup>2</sup> | Overall |  | Average |  | Below average |  | Above average |  |
| --- | --- | --- | --- | --- | --- | --- | --- | --- | --- |
|  |  | Accuracy<sup>3</sup> | Kappa<sup>4</sup> | Sensitivity<sup>5</sup> | Specificity<sup>6</sup> | Sensitivity | Specificity | Sensitivity | Specificity |
| 1. boot_RFE | 13 | 0.61 | 0.39 | 0.60 | 0.64 | 0.74 | 0.85 | 0.46 | 0.89 |
| **2. boot_RFE_selected** | 8 | **0.63** | **0.42** | **0.60** | **0.67** | **0.74** | **0.85** | **0.54** | **0.89** |
| 3. CV_RFE | 13 | 0.60 | 0.38 | 0.52 | 0.69 | 0.77 | 0.81 | 0.54 | 0.87 |
| 4. CV_RFE_selected | 8 | 0.56 | 0.32 | 0.52 | 0.62 | 0.64 | 0.82 | 0.54 | 0.86 |
| 5. boot_LASSO | 29 | 0.61 | 0.39 | 0.56 | 0.67 | 0.77 | 0.83 | 0.50 | 0.87 |
| 6. boot_EN | 67 | 0.58 | 0.34 | 0.56 | 0.62 | 0.74 | 0.82 | 0.42 | 0.89 |
| 7. CV_LASSO | 29 | 0.58 | 0.35 | 0.52 | 0.65 | 0.81 | 0.82 | 0.42 | 0.86 |
| 8. CV_EN | 67 | 0.55 | 0.31 | 0.48 | 0.64 | 0.77 | 0.79 | 0.42 | 0.86 |

Note: 1. 'boot' refers to models using bootstrap aggregating as the sampling method; 'CV' refers to models using cross-validation as the sampling method; 'RFE' refers to features selected using the recursive feature elimination approach; 'RFE\_selected' refers to the RFE features which were further selected by removing correlated features and features not related to SIR; LASSO' refers to features selected using the least absolute shrinkage and selection operator approach; 'EN' refers to features selected using the elastic net approach.
2. Lists of predictors for model 1-4 were presented in Appendix 10.
3. Accuracy refers to the proportion of predictions that were correctly predicted.
4. Kappa is a measure of agreement between actual and predicted classes, considering the agreement occurring by chance.
5. Sensitivity refers to the proportion of positives that were true positives.
6. Specificity refers to the proportion of negatives that were true negatives.

In addition, the performance of LcVI in predicting the SIR of lung cancer was compared with the performance of the Australian area disadvantage index, the Relative Socio-economic Disadvantage (IRSD) index [43]. The IRSD index is a general socio-economic index that summarises a range of information about the economic and social conditions of people and households within an area. It ranges from one to ten, with a score of one representing the least disadvantaged areas and ten representing the most disadvantaged areas. The same statistical tests and visualisations were also used to evaluate the capacity of the IRSD to explain the geographic variation of lung cancer incidence in Queensland.

All analyses were performed with R version 4.4.0. Bayesian spatial model was fitted using the ‘*CARBayes’* package (version 6.1.1) and random forest models used the ‘*caret’* package (version 6.0.94). Appendix 9 gives an example of syntax showing the key functions used in the REWIRED framework.

## Results

Between 2016 and 2019, the SIR of lung cancer for the 519 SA2s ranged from 0.46 to 1.60. There was evidence that lung cancer incidence rates were below the Queensland average in 158 (30.4%) SA2s, above the average in 120 (23.2%) SA2s, and not different from the average in the remaining 241 (46.4%) SA2s.

Eight random forest models were developed based on different variable selection and sampling methods (Table 1). Model 2 based on eight predictors selected using the RFE method (RFE_selected predictors) and bagging sampling method achieved the highest Kappa statistics (Kappa = 0.42) and accuracy (accuracy = 0.63, 95% CI: 0.53-0.72, no information rate=0.47, p<0.001) in the test set (See Appendix 6&11 for more information). The model correctly identified 74% of the ‘below average’ SA2s, 54% of the ‘above average’ SA2s, and 60% of the ‘average’ SA2s, indicating its sensitivity for each category. In terms of specificity, the model correctly identified 85%, 89%, and 67% of areas as not belonging to the ‘below average,’ ‘above average,’ and ‘average’ categories, respectively.

Table 2 lists the eight RFE_selected predictors with their feature important scores generated by Model 2. The prevalence of diabetes had the highest feature importance score. SA2s categorised as ‘below average’ had a median age-standardised prevalence rate (ASR) of diabetes as 3.78, contrasting with a median ASR of 6.07 for SA2s with ‘above average’ lung cancer incidence. The univariate linear regression model showed that this variable alone explained 45% of the lung cancer geographic variation (R^2^=0.45, Table 2).

**Table 2.** Feature importance scores and median values^1^ by categories of lung cancer incidence for the selected predictors.

| Features | Feature importance score | Median values by category |  |  | R square<sup>3</sup> |
| --- | --- | --- | --- | --- | --- |
|  |  | Below | Average | Above |  |
| Diabetes mellitus (ASR<sup>2</sup>) | 81.55 | 3.78 | 4.77 | 6.07 | 0.45 |
| Adequate fruit intake (ASR) | 58.35 | 52.71 | 50.52 | 48.59 | 0.33 |
| High-income taxpayers with private health insurance (%) | 55.69 | 56.49 | 46.45 | 33.09 | 0.37 |
| Bachelor’s degree (%) | 48.19 | 18.25 | 10.30 | 7.25 | 0.43 |
| Unemployed (%) | 44.52 | 5.90 | 6.90 | 10.00 | 0.18 |
| Median annual income-Superannuation and annuity (\$1000) | 34.58 | 20.52 | 17.27 | 15.09 | 0.24 |
| Highest year of school completed-Year 11 or equivalent (%) | 32.63 | 6.60 | 8.10 | 8.60 | 0.28 |
| Marital status-Separated (%) | 29.73 | 2.90 | 3.60 | 4.35 | 0.29 |

*Note: 1. Values were standardised when included in random forest models; 2. ASR: age standardised prevalence rate per 100 people. 3. $R^2$ is based on linear regression between continuous $\log_2(\text{SIR})$ and the specific feature.*

Based on the median values, the prevalence rates of diabetes, being unemployed, lower education level, and a marital status of ‘separated’ were positively associated with lung cancer incidence; while the prevalence rates of adequate fruit intake, having private health insurance and higher education, and the median superannuation and annuity income were negatively associated with lung cancer incidence.

Areas with ‘below average’ SIR had the lowest scores of the generated index (LcVI) (median=2.28, Q1-Q3: 0.85-3.72) compared to SA2s with ‘average’ SIR (median LcVI=5.30, Q1-Q3: 3.88-6.55) and ‘above average’ SIR (median LcVI=8.03, Q1-Q3: 6.98-9.16) (Figure 2). The ANOVA test showed significant differences in the LcVI values across the three categories of SIR (F (2, 516) = 287.8, p<0.001). Post-hoc analysis further suggested a clear gradient in the LcVI by SIR, with the ‘below average’ category having a significantly lower LcVI compared to the ‘average’ (mean difference = 2.80, 95% CI: 2.34-3.25, p < 0.001); while the ‘above’ category had a significantly higher LcVI than the ‘average’ (mean difference = 2.70, 95% CI: 2.20-3.19, p < 0.001).

**Figure 2.**
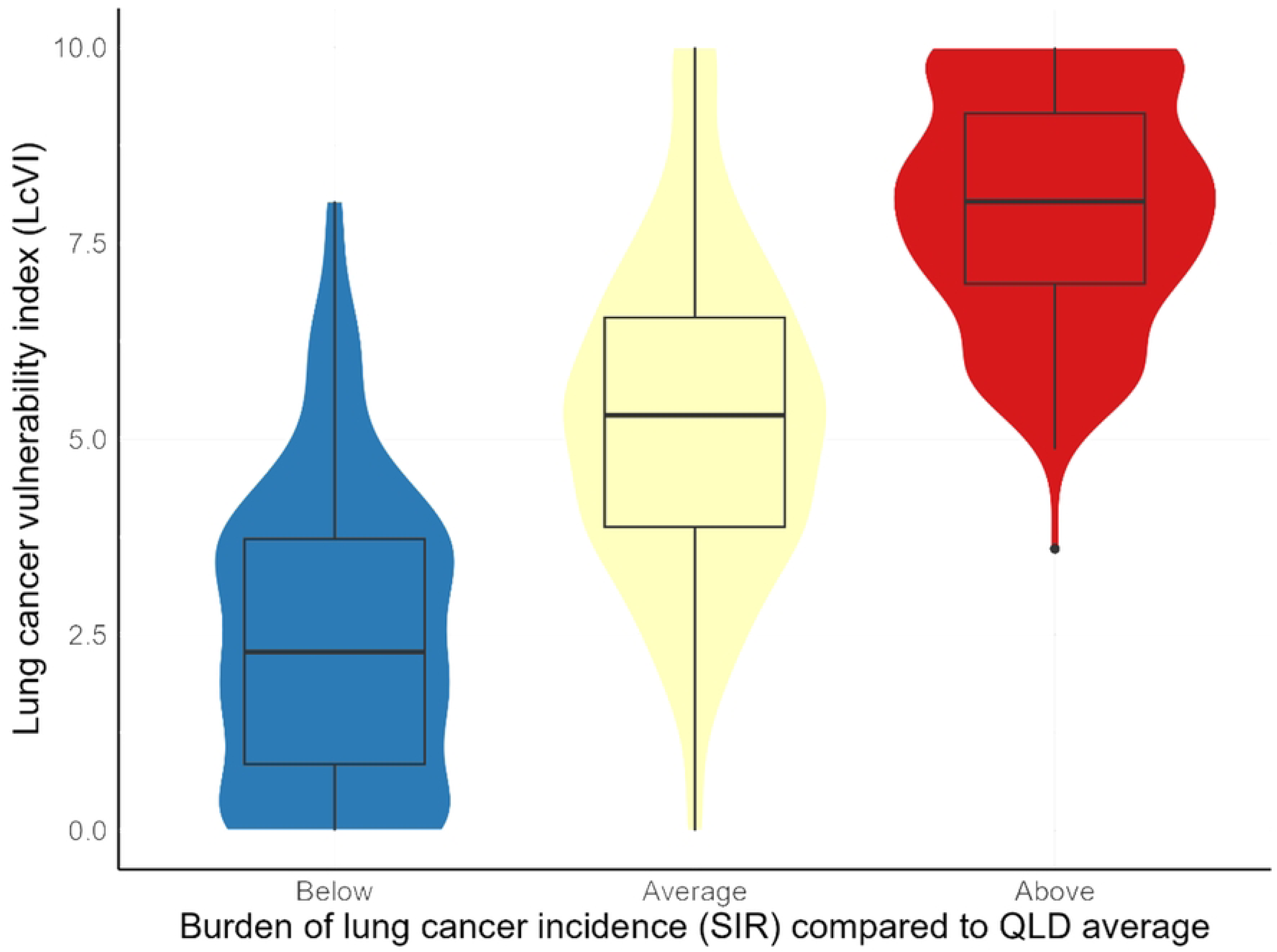
Violin plot with boxplot of lung cancer vulnerability index (LcVI) by categories of lung cancer standardised incidence ratio (SIR).

The LcVI was significantly associated with the continuous SIR for the 519 SA2s, with a Peason correlation of 0.76. On linear regression, the LcVI explained 57% of the variation in the (natural) logarithm-transformed SIR across Queensland, with each one-point increase in LcVI associated with a 6% increase in SIR (R^2^=0.57, coef.=0.06, t=26.3, p<0.001). In the top graph of Figure 3, the triangles and dots for each column represent the LcVI and SIR of the same SA2, with colour representing the value of SIR. Most of the SA2s with low SIR (blue) had low LcVI (triangles down in the bottom), and vice versa. The comparative analysis showed that, compared to the IRSD index, the LcVI explained more variation in SIR across Queensland (42% vs 57%). The lower graph of Figure 3 also shows a less significant association between SIR and IRSD.

**Figure 3.**
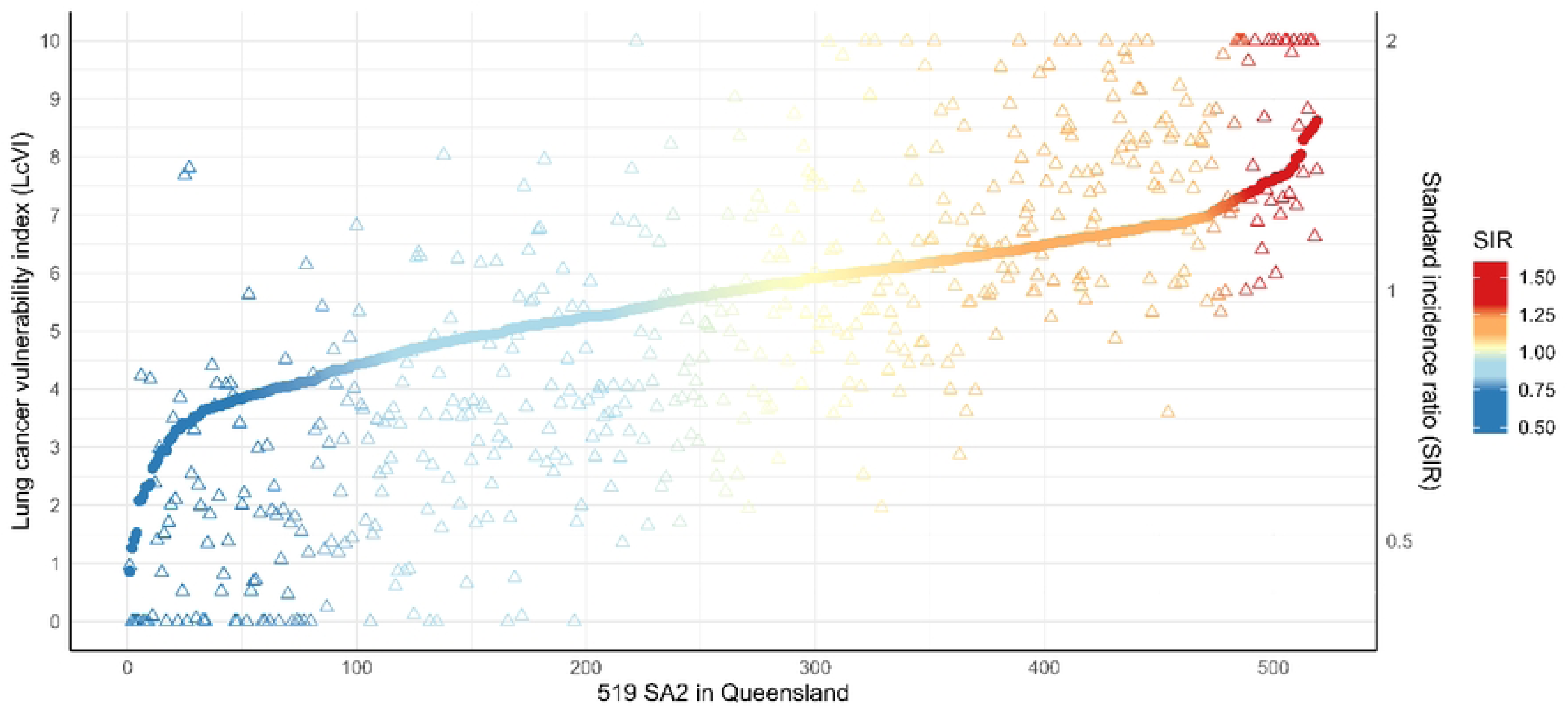
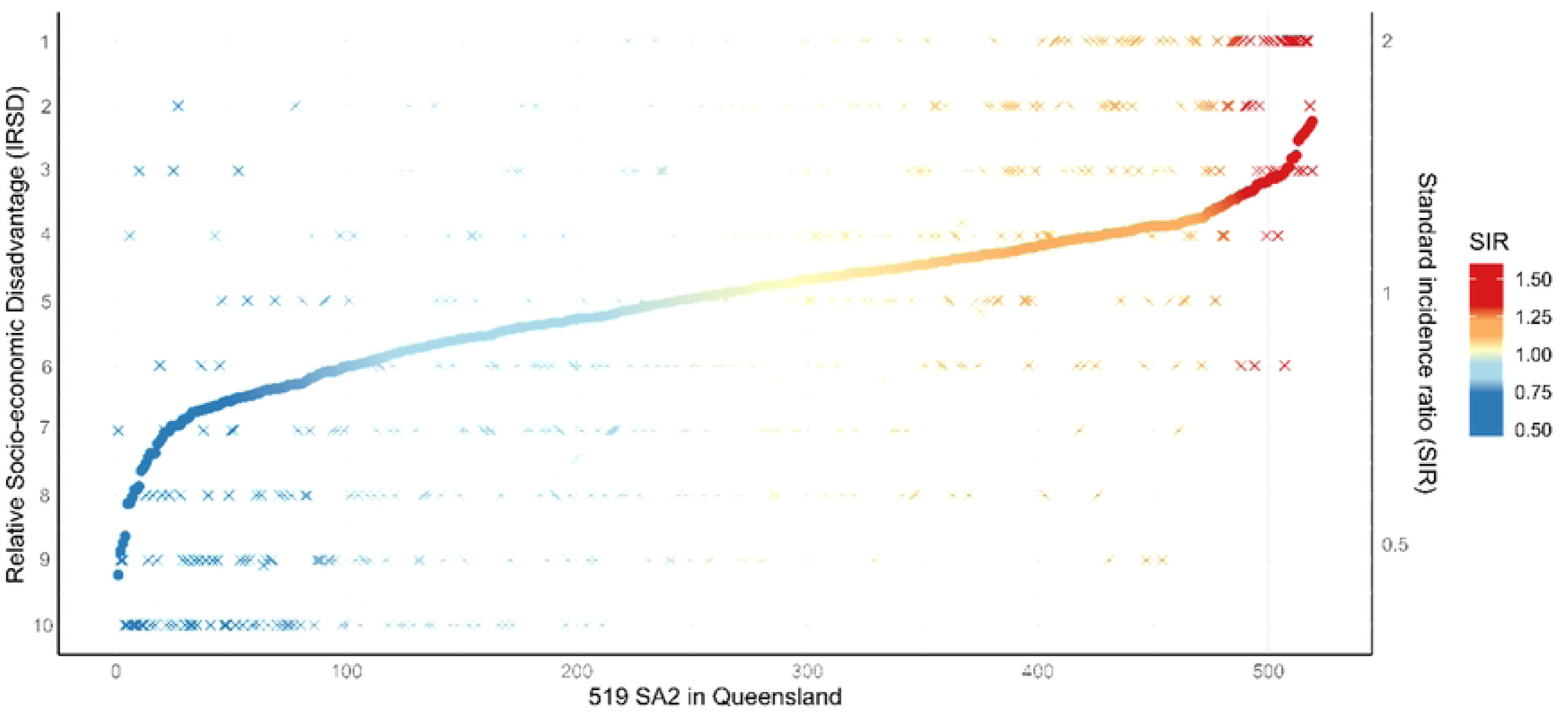
Values of lung cancer vulnerability index (LcVI, upper, triangles) or relative socio-economic disadvantage (IRSD, lower, crosses) and lung cancer standardised incidence ratio (SIR, dot) for the 519 SA2s in Queensland. Note: The y-axis for IRSD has been reversed to make the trend of the IRSD plot consistent with the trend of the SIR plot; The colours of triangles, crosses and dots represent the values of SIR. The order that the SA2s appear on the x-axis is arranged by SIR values.

## Discussion

Lung cancer is one of the most preventable cancers, with over 80% of cases attributable to tobacco smoking and other environmental risk factors [44]. However, due to the significant latency period between exposure to these risk factors and lung cancer diagnosis [45], it is difficult to establish a direct relationship between the prevalence of risk factors and lung cancer incidence in ecological studies. While not specifically overcoming this limitation, this study proposes a standard framework, the REWIRED approach, to identify important area-level characteristics that might serve as a proxy to reflect the historical prevalence of lung cancer risk factors, thus providing further understanding of reasons for the geographic variations of lung cancer incidence. Based on eight area-level characteristics, the final model achieved high specificity (0.89) for predicting areas with above-average incidence. This high specificity indicates that if the model predicts an area as not above average, it is highly likely that the lung cancer incidence in that area is indeed not above average. Similarly, the model performs well in predicting areas with below-average incidence, achieving a specificity of 0.85, meaning it effectively avoids falsely identifying areas as below average when they are not. The lung cancer specific index generated based on the eight predictors explained more (57% vs 42%) of the geographic variation compared to the more generic area-level socioeconomic disadvantage index based on 16 characteristics [43].

There are several advantages of using the REWIRED approach. Firstly, this approach is flexible with different data patterns by using non-parametric techniques in both supervised variable selection (Random Forest models) and dimensional reduction (gap-adjusted percentile rank-sum) processes. Compared with regression models for feature selection and PCA for dimensional reduction, this non-parametric approach is able to address non-linear data, which is a typical characteristic of area-level datasets. Comparative analysis also showed that the non-parametric variable selection using RFE performed better than the parametric variable selection methods using LASSO or EN methods.

In contrast to common unsupervised dimensional reduction approaches such as PCA and percentile rank sum which treat all variables equally without considering their relationship with health outcomes, the REWIRED approach weighted the predictors based on the feature importance scores derived from Random Forest models, which allowed for the construction of a vulnerability index that reflects the relative importance of each variable.

To our knowledge, our study is the first ecological study to report a link between the population prevalence of diabetes and the incidence of lung cancer. The age-standardized prevalence rate of diabetes achieved the highest feature importance during the REWIRED process, explaining 45% of the geographic variation in lung cancer incidence. Previous studies have linked increased diabetes risk with various smoking behaviours, including active smoking, passive smoking, and smoking cessation, showing a dose-response relationship between smoking and diabetes risk [46]. Therefore, while our results would need confirmation through other studies, the population prevalence of diabetes could potentially provide a more effective indicator of long-term smoking behaviours among the residents compared to the current smoking prevalence. In addition, since diabetes usually causes symptoms and requires treatment, population-level estimates of diabetes prevalence could be more reliable and accessible than corresponding estimates of smoking prevalence. Our findings showed a high correlation between the prevalences of diabetes and current smoking at the small area level (Pearson’s correlation coefficient = 0.78). Of note, the predictor relating to current smoking prevalence had a lower feature importance score in predicting lung cancer incidence so was excluded from the final model. This could be expected given the often 20-30 year lag period between smoking patterns and lung cancer incidence patterns [47]. This may increase the rationale for using diabetes prevalence as an ecological measure of previous smoking behaviour, rather than current smoking prevalence.

The prevalence of adequate fruit intake was identified as the second most important feature related to the spatial variation of lung cancer incidence in our study. Inadequate fruit intake has previously been identified as having a causal association with lung cancer, and is an attributable factor in as many as 9.6% of lung cancer diagnoses [44, 48]. In addition, a meta-analysis of 38 individual-level studies also reported a significant protective effect of adequate fruit intake against lung cancer [49]. On the area level, a more plausible explanation for our results may be that adequate fruit intake is a proxy for broader health-conscious behaviours, as individuals who meet dietary guidelines for fruit consumption are likely to engage in other healthy lifestyle practices [50]. Additionally, the Australian Cancer Atlas 2.0 indicates similarity between those areas that have a higher prevalence of health risk factors such as smoking and those areas that have higher rates of inadequate food intake, further supporting the suggested link between dietary habits and overall health behaviours [2].

It is well-established that the population living in socioeconomically disadvantaged areas tend to have higher lung cancer incidence rates [5–7]. Our study identified six key socioeconomic features related to lung cancer risk, providing further insight into the association between area disadvantage and lung cancer. Some of these features overlap with those used in generating the IRSD index [43]. Whether these features are individually important in explaining the variation in lung cancer diagnosis, or collectively measure some aspect of socioeconomic status, is not known.

## Conclusion

This study developed a novel framework to conduct robust ecological analyses of lung cancer outcomes, with the potential to extend this to other cancer types and outcomes. The REWIRED approach showed better explanatory power for geographic variations in lung cancer incidence compared to standard population-based area-level socioeconomic indices. While these area-level results cannot be extrapolated to the individual level, by using easily accessible ecological data they provide important insights into the underlying drivers of the observed geographic disparities in lung cancer incidence and so have an important role in guiding further research.

## Data Availability

The links for area-level variables have been provided in references. Modelled area-level lung cancer incidence rates are available by contacting the corresponding author.
Australian Urban Research Infrastructure Network. ABS - Data by Region - Economy & Industry (SA2) 2011-2019. 2023 [cited 2024 Available from: https://data.aurin.org.au/dataset/au-govt-abs-abs-data-by-region-economy-and-industry-asgs-sa2-2011-2019-sa2-2016. 20. Australian Urban Research Infrastructure Network. ABS - Data by Region - Population & People (SA2) 2011-2019. 2023 Available from: https://data.aurin.org.au/dataset/au-govt-abs-abs-data-by-region-pop-and-people-asgs-sa2-2011-2019-sa2-2016. 21. Australian Urban Research Infrastructure Network. ABS - Data by Region - Income (Including Government Allowances) (SA2) 2011-2019. 2023 [cited 2024 Available from: https://data.aurin.org.au/dataset/au-govt-abs-abs-data-by-region-income-asgs-sa2-2011-2019-sa2-2016. 22. Australian Urban Research Infrastructure Network. ABS - Data by Region - Health & Disability (SA2) 2011-2018. 2023 [cited 2024 Available from: https://data.aurin.org.au/dataset/au-govt-abs-abs-data-by-region-health-and-disability-asgs-sa2-2011-2018-sa2-2016. 23. Australian Urban Research Infrastructure Network. ABS - Data by Region - Family & Community (SA2) 2011-2018. 2023 [cited 2024 Available from: https://data.aurin.org.au/dataset/au-govt-abs-abs-data-by-region-family-and-community-asgs-sa2-2011-2018-sa2-2016. 24. Australian Urban Research Infrastructure Network. ABS - Data by Region - Education & Employment (SA2) 2011-2019. 2023 [cited 2024 Available from: https://data.aurin.org.au/dataset/au-govt-abs-abs-data-by-region-education-and-employment-asgs-sa2-2011-2019-sa2-2016. 25. Public Health Information Development Unit. About PHIDU. [cited 2024 Available from: https://phidu.torrens.edu.au/about-phidu. 26. Australian Urban Research Infrastructure Network. PHIDU - Prevalence of Selected Health Risk Factors - Adults (PHA) 2017-2018. 2023 [cited 2024 Available from: https://data.aurin.org.au/dataset/tua-phidu-phidu-estimates-risk-factors-adults-pha-2017-18-pha2016. 27. Australian Urban Research Infrastructure Network. PHIDU - Prevalence of Chronic Diseases (PHA) 2017-2018. 2023 [cited 2024 Available from: https://data.aurin.org.au/dataset/tua-phidu-phidu-estimates-chronic-disease-pha-2017-18-pha2016.

## Acknowledgments

The authors express their gratitude to the Data Custodian, as well as the staff of the Australian Urban Research Infrastructure Network (AURIN) and the Queensland Cancer Register (QCR), for their support in providing the relevant datasets.

## Supporting information captions

Appendix 1 Modelling of indirect standardized incidence ratios

Appendix 2 List of area-level variables

Appendix 4 Number of health facilities within each category in Queensland

Appendix 5 The density of health services in each SA2 area

Appendix 6 Introduction of model evaluation statistics

Appendix 7 Example of gap adjusted percentile ranking.

Appendix 8 Feature selection using LASSO and EN

Appendix 9 Example of syntax for REWIRED framework

Appendix 10 Lists of predictors included in model 1-4.

Appendix 11 Confusion matrix based on test set predicted by Model 2

## Notes

### Competing Interest Statement

The authors have declared no competing interest.

### Funding Statement

The author(s) received no specific funding for this work.

### Author Declarations

Approval was obtained from the data custodian to access de-identified routinely collected cancer incidence data from Queensland Cancer Register (QCR).

